# Non-intuitive trends of fetal fraction development related to gestational age and fetal gender, and their practical implications for non-invasive prenatal testing

**DOI:** 10.1101/2022.04.07.22273556

**Authors:** Natalia Forgacova, Juraj Gazdarica, Jaroslav Budis, Marcel Kucharik, Martina Sekelska, Tomas Szemes

## Abstract

**Objective:** Discovery of fetal cell-free DNA fragments in maternal blood revolutionized prenatal diagnostics. Although non-invasive prenatal testing (NIPT) is already a matured screening test with high specificity and sensitivity, the accurate estimation of the proportion of fetal fragments, called fetal fraction, is crucial to avoid false-negative results.

**Methods:** We collected 6999 samples from women undergoing NIPT testing with a single male fetus to demonstrate the influence of fetal fraction by the maternal and fetal characteristics.

**Results:** We show several fetal fraction discrepancies that contradict the generally presented conventional view. At first, the fetal fraction is not consistently rising with the maturity of the fetus due to a drop in 15 weeks of maturation. Secondly, the male samples have a lower fetal fraction than female fetuses, arguably due to the smaller gonosomal chromosomes. Finally, we discuss not only the possible reasons why this inconsistency exists but we also outline why these differences have not yet been identified and published.

**Conclusion:** We demonstrate two non-intuitive trends to better comprehend the fetal fraction development and more precise selection of patients with sufficient fetal fraction for accurate testing.

**Bulleted statements:** *What is already known about this topic?:*

- Non-invasive prenatal testing has become a well-known mature screening test, and the fetal fraction is studied in detail by research teams worldwide.

*What does this study add?:*

- Here we demonstrate two non-intuitive trends to better comprehend fetal fraction development that can further increase the sensitivity of routine testing by proper selection of blood sampling according to gestational age and fetus gender.

## INTRODUCTION

Prenatal diagnosis plays an important role in current gynecological practice and primary health care for pregnant women. Initially, prenatal diagnostic methods relied on invasive techniques to obtain a sample of fetal biological tissue. However, these methods represent certain risks for the developing fetus and the pregnant woman ^1^. New hope for non-invasive prenatal diagnosis appeared in 1997, discovering cell-free fetal DNA (cffDNA) in maternal plasma and serum samples ^2^. Since then, cffDNA has become a unique genetic material for prenatal screening. In no other medicine area has extracellular DNA been introduced into clinical practice as rapidly as cffDNA and non-invasive prenatal testing (NIPT) ^3^. Based on the analysis of cffDNA in maternal plasma by massively parallel sequencing (MPS), NIPT has become increasingly popular and is currently represented as the most sensitive and effective prenatal screening tool for detecting common fetal chromosomal aneuploidies such as trisomy 21, 18, and 13 ^4^.

The circulating cell-free DNA (cfDNA) in maternal blood is a mixture of predominant genomic DNA fragments released from the mother’s hematopoietic system and fetal DNA derived from apoptotic trophoblastic cells in the placenta. The amount of fetal cfDNA compared to the total cfDNA in maternal circulation, known as the fetal fraction (FF), is an important critical parameter for ensuring NIPT’s accuracy and interpreting the clinical outcomes ^5,6^. Previous studies suggested that the average FF is 10-15% between 10 and 20 weeks of gestation; however, FF is a dynamic value that can vary greatly between individual pregnancies and throughout the pregnancy ^7,8^. Although most NIPS tests are successful, 1-8% of pregnant women has uninformative, non-reportable, or no-call results ^9^. There are essentially two main reasons for failing to achieve a result. First, technical problems with blood collection, transportation of the samples, storage, or problems with laboratory processing, including failed DNA extraction, amplification, and sequencing ^10^. Second, the most common reason for test failure is insufficient FF (the minimum FF threshold varies by assay, but typically around 4%) ^9^. In recent years, a number of maternal and fetal characteristics have been identified that affect FF.

Both FF and the odds of receiving a result on a first draw have been shown to decrease with maternal weight and increase with gestational age ^11–14^. It is well known that FF increases with later gestational age ^15,16^. However, the rate of increase in FF is not constant across different gestational ages. In general, the lowest rate of increase with gestational age occurs between 10 and 21 weeks of gestation. Starting at 21 weeks of gestation, a noticeably greater weekly increase in FF is observed and remains constant from then onwards ^5,17,18^. There have also been reports of a temporary decrease in FF from the first to the second trimester due to increased maternal weight during this gestational time ^19^.

While FF is considered a key parameter to obtaining accurate NIPT results, factors that affect FF have not previously been studied in the Slovak population. The objective of this study was to compare FF in different gestational ages and examine if a relationship exists between a decrease in FF and an increasing maternal weight at a certain gestational age. We draw comparisons with previous studies and provide further insight into these complex relationships. We also compare FF between fetal gender to add new evidence to the understanding of factors affecting FF, which is also important for the clinical application of NIPT.

## METHODS

### Sample Acquisition

We collected 6999 samples from women undergoing NIPT testing with a single male fetus, from which 6536 samples are negative (healthy), and 463 samples are uninformative (due to the low FF), respectively. The FF, according to the method based on the proportion of fragments on the Y chromosome ^15^, has been calculated for these samples. Our work was part of two clinical studies approved by the Ethical Committee of the Bratislava Self-Governing Region (Sabinovska ul.16, 820 05 Bratislava): the first one called “NIPT study” (study ID 35900_2015 approved on 30 April of 2015 under the decision ID 03899_2015) and the second one called “SNiPT” (study ID 37136/2018 approved on 11 June of 2018 under the decision ID 07507/2018/HF). All patients in the study signed written informed consents consistent with the Helsinki declaration, which the ethics mentioned above committee approved.

More detailed information about the NIPT dataset generated and analyzed in this study is fully described in our previous studies ^20,21^.

### Data Preparation and Evaluation of Models

FFs were calculated using gender-independent methods based on the combination of the FF predictors based on the independent sources of information (Combo method) ^22^. FFs were also calculated with the reference Y-based method described below.

### Y-Based Estimator

FF based on the proportion of the Y was determined according to equation ^15^,

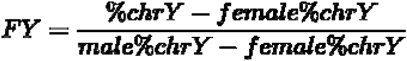

where male *%chrY* and female *%chrY* is the mean fraction of chromosome Y sequence reads of plasma samples obtained from 14 adult male individuals and pregnant women bearing euploid female fetuses, respectively. *%chrY* is the fraction of chromosome Y sequence reads of the sample whose FF we want to calculate.

## RESULTS

### Sample characteristics

A total of 6999 women with singleton pregnancies undergoing NIPS were used for the present study. The basic pregnancy characteristics of the samples (6999), including maternal age, weight, height, body mass index (BMI), gestational age, and FF, are presented in Table 1. The minimum gestational age was only 11 weeks, and the maximum was 24 weeks, with a median gestational age at the time of testing of 14 weeks (Figure 1A). When considering the FFweeks, data ranged from 1.1% to 44.9%, with a median of 11% (Figure 1B). The maternal age ranged from 15 to 52 years old, with a median of 35 years old (Figure 1C). Considering the BMI, the lowest and highest BMI of subjects in the group was found to be 13.8 kg/m2 and 47.7 kg/m2, with a median of 23.4 kg/m2 (Figure 1D).

**Table 1.** Maternal and fetal characteristics of the study population (n=6999).

| **Characteristic** | **Mean** | **Median** | **Std.**<br>**deviation** | **Minimum** | **Maximum** |
| --- | --- | --- | --- | --- | --- |
| **Maternal age**<br>**(years)** | 34 | 35 | 4.9 | 15 | 52 |
| **Weight (kg)** | 68.9 | 66 | 14.1 | 36 | 151 |
| **Height (cm)** | 167.5 | 168 | 6.1 | 131 | 197 |
| **Body mass**<br>**index (kg/m2)** | 24.5 | 23.4 | 4.8 | 13.8 | 47.7 |
| **Gestational**<br>**age (weeks)** | 14.5 | 14 | 2.6 | 11 | 24 |
| **Fetal fraction**<br>**(%)** | 11.6 | 11 | 4.9 | 1.1 | 44.9 |

**Figure 1.**
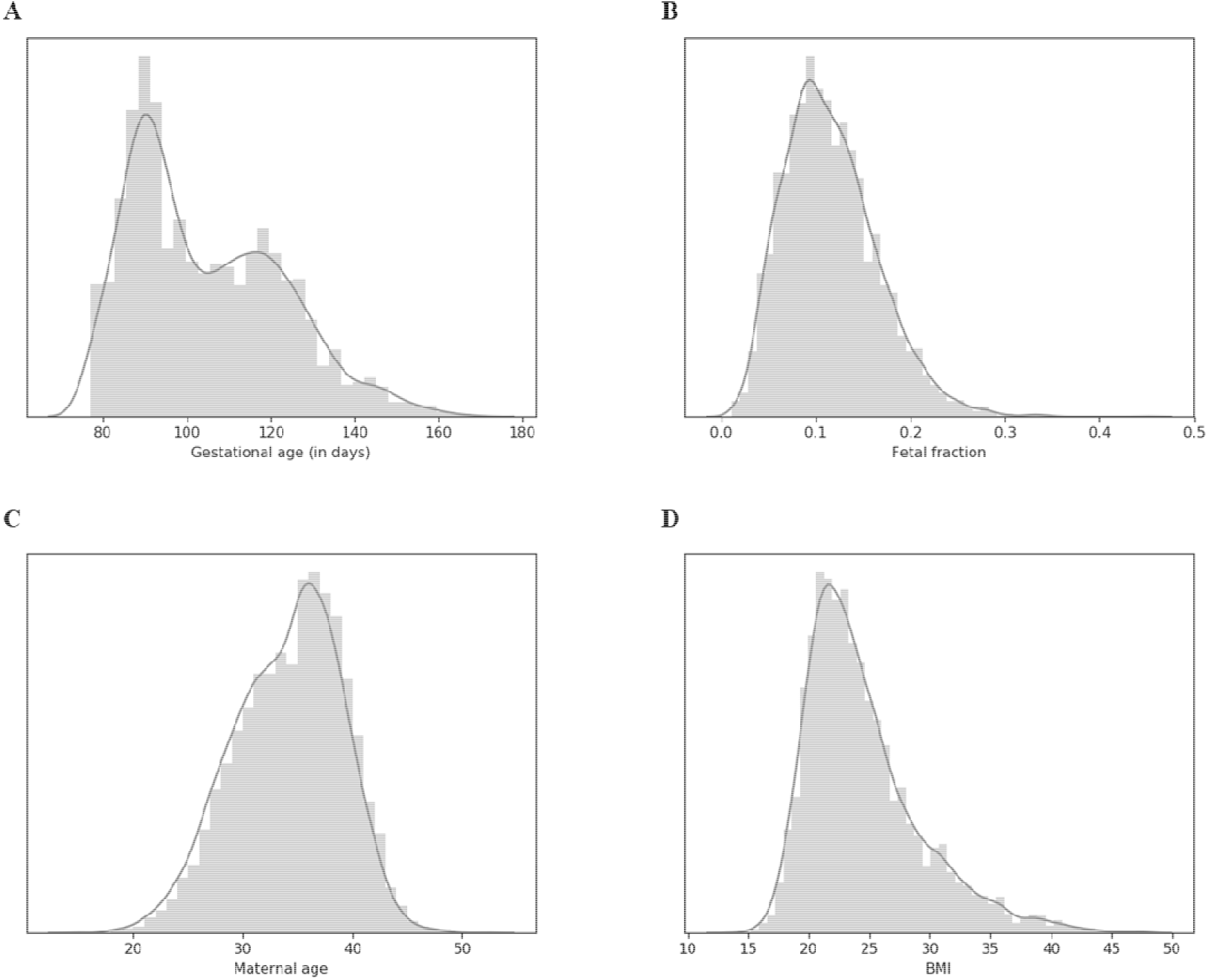
Distributions of gestational age (A), fetal fraction (B), maternal age (C) and body mass index (D) across all samples in this study (n=6999).

### Effect of gestational age on the fetal fraction

In agreement with previous studies, our results reveal that FF is positively correlated with gestational age. However, overall the rate of increase in FF is not constant across different gestational ages. From 11 to 23 weeks’ gestation, we detected only a slight increase in the median value for the FF (Figure 2A). The rate of increase becomes noticeably higher by around 24 weeks gestation. On average, the FF at around 24 weeks gestation onwards (approximately 20%) is more than twice that observed at up to 23 weeks gestation (approximately 10%) (Figure 2A). However, further examination using distribution FF in days showed one interesting trend that provides closer sight of the effect of FF during different gestational ages. From 77 days’ gestation (approximately 11 weeks gestation), median FF reached a minimum value of 10.44%. Over this day, FF noticeably increased to the median FF value of 11.34% in 92 days’ gestation (approximately 13 weeks gestation). Median FF at this point corresponds to median FF for the 20 weeks’ gestation. From 92 days’ gestation, we observed a significant decrease in FF to 10.8% in 102 days’ gestation (approximately 14 weeks gestation). After this critical point, median FF continued to increase again, as between 17 to 20 weeks’ gestation remained constant and in 168 days’ gestation reached the median FF of 13.11% (Figure 2B). To verify the probable cause of this decrease in FF, we demonstrated the relationship between FF and maternal weight in different gestational ages. The results show a sharp increase in maternal weight in the time interval where FF decreases significantly (Figure 2B). We also illustrated a loess-smoothed line for FF in different gestational days in male samples (Figure 2C).

**Figure 2.**
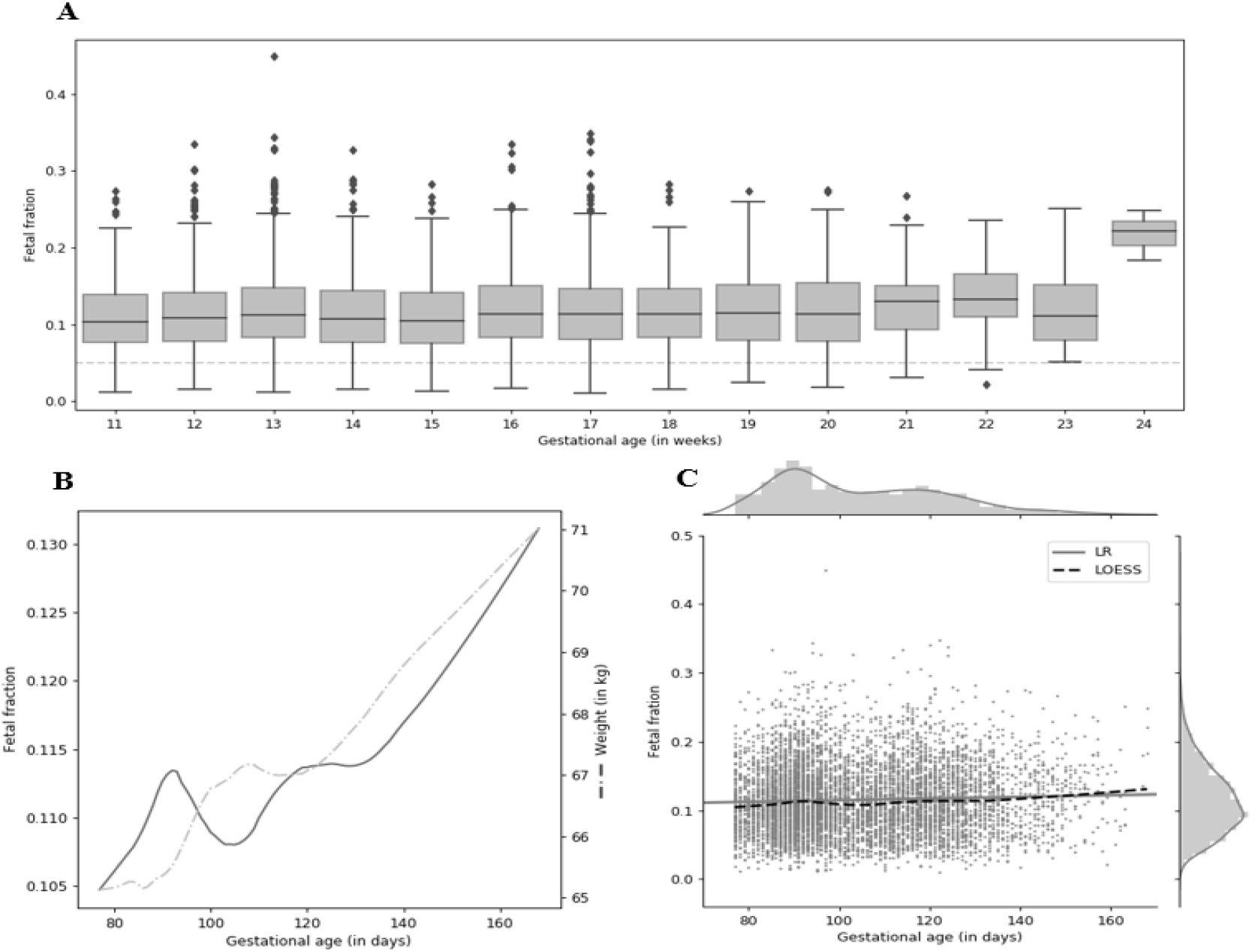
Effect of gestational age on the fetal fraction. (A) The effect of gestational age on the percentage of FF in different gestational weeks. (B) Shows Loess-smoothed trends for FF and weight in male samples. At day 102 of gestational age, which corresponds to 15 week of gestational age (between days 99-105 of gestational age) we found a local minimum for median FF calculated for each day of sampling. (C) Loess-smoothed line for FF in different gestational days in male samples.

### Effect of fetal gender on the fetal fraction

We examined the relationship between fetal gender and FF. We used the Y-based ^15^ method to calculate the FF for male samples and the gender-independent Combo method ^22^ to calculate the FF for male and female samples. This method combines the state-of-the-art SeqFF method ^23^ and the method estimating FF based on fragment length. Comparing the results, we found statistical significance for comparing FF between observed groups - Y based, Combo XY, and Combo XX (Figure 3A) (p-value = 1.35e-12 from Kruskal-Wallis test). For the male samples, the Y-based method corresponded to the Combo method (p-value = 0.44 from Mann Whitney U test), but for female samples, the FF values were significantly higher for the Combo method compared to the Combo method for male samples (by 0.6%, p-value = 1.79e-10 from Mann Whitney U test) and also for Y-based method (by 0.6%, p-value = 6.25e-12 from Mann Whitney U test) (Figure 3A). Since the Combo method is based on the fragment lengths profiles combined with the SeqFF method, we studied the differences between genders’ median fragment length. Figure 3B shows concordance for the median male and female fragment length distribution (p-value = 1 from Kolmogorov-Smirnov test). However, by highlighting the differences by subtracting male and female fragment length profiles, we found that the shorter fragments preferably refer to female samples (to 150 bp) and longer fragments to male samples (Figure 3C). This result corresponds to the finding that the higher the proportion of shorter fragments is, the higher FF is observed ^22^.

**Figure 3.**
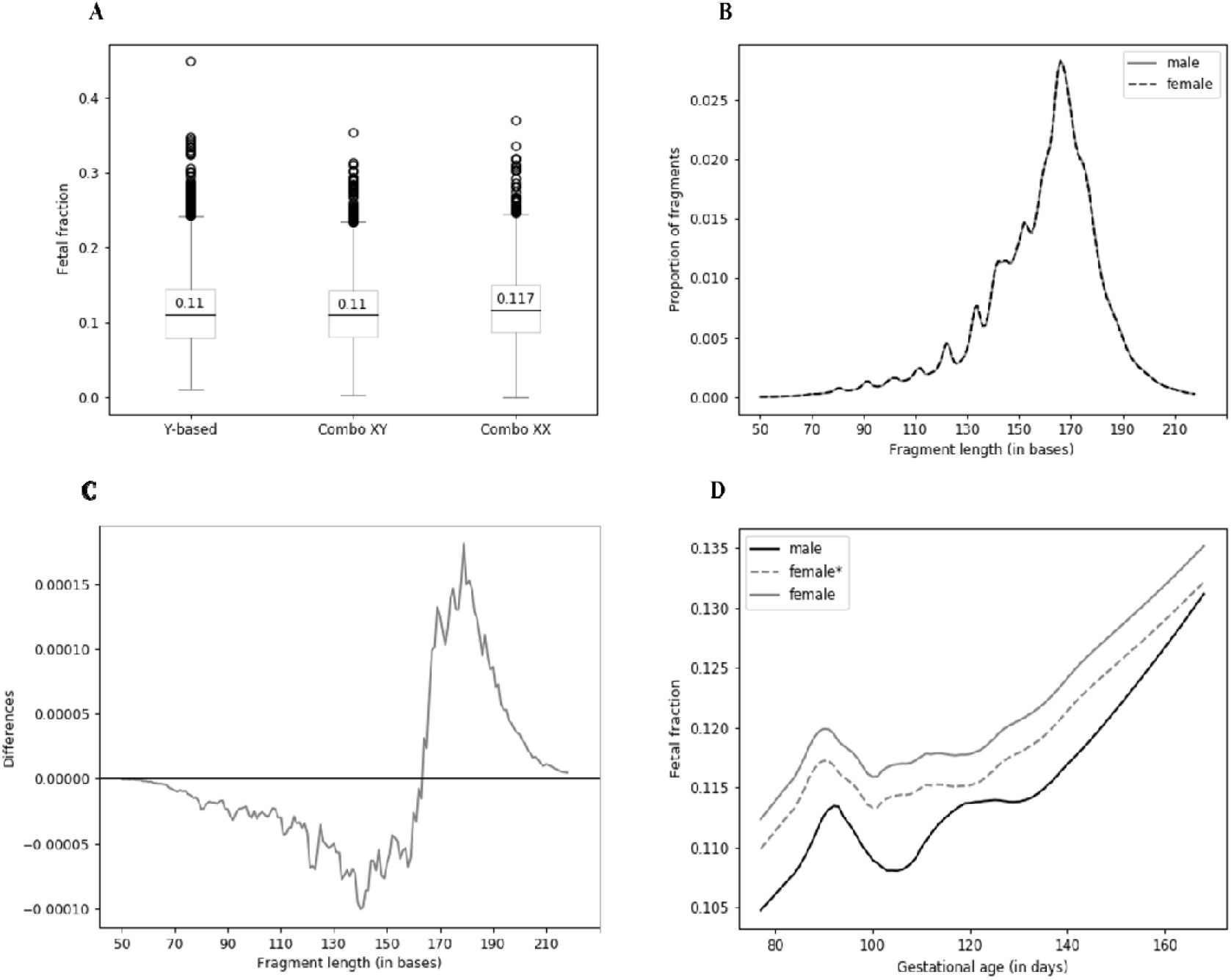
Effect of fetal gender on the fetal fraction. (A) Comparison of FF quantification by Y-based method (male samples) and Combo XY and XX methods for male and female samples, respectively. (B) Median fragment-length profiles for male and female samples (p-value = 1 from Kolmogorov-Smirnov test). (C) Comparison of the fragment length profiles between male and female samples (grey curve represents subtraction of male and female median length profiles, respectively). (D) Comparison of male and female fetal fraction profiles based on gestational age. Compensation for the longer genome is displayed as dashed line (female*).

This inconsistency between FFs with different lengths of male and female genomes can be partially explained by the uneven lengths of sex chromosomes between fetal genders. Since the X chromosome is much longer than the Y chromosome, the female genome is overall slightly longer than the male genome, and thus the female fetuses will release a bit more cfDNA into the maternal bloodstream than the male fetuses. To evaluate the extent of this effect, we compared what portion of all mapped reads are mapped to autosomes. In our setting, the ratio of autosomes is 97.43% for non-pregnant male samples and 95.27% for non-pregnant female samples, so 1.0227 times more reads were mapped to autosomes in a male sample. Since the length of autosomes is equal for male and female individuals, the estimated mappable length of female genomes is longer by 2.27% than the male genome. Note that these numbers can vary for different used protocols and mapping software.

The FF estimation is linearly dependent on the number of mapped reads from fetuses; thus, to compensate, we divided the female FF by this number (1.0227). However, this compensation explained only slightly less than half of the differences between male and female FFs that we see (Figure 3D).

## DISCUSSION

Although the NIPT is a screening test with high specificity and sensitivity, a small percentage of tests fail due to low fetal FF. The factors that influence FF have been of interest to various researchers over the years. Some biological factors are well-known to affect FF, such as maternal weight and gestational age. On the contrary, the FF difference between fetal gender and the effect of other maternal characteristics is still under investigation. Our study focused on a cohort of 6999 pregnant women with singleton pregnancies undergoing NIPT in Slovakia. We identified 463 cases in which an initial sampling yielded a failed result due to low FF. Our results showed that the FF of female fetuses was significantly higher than male fetuses at the same gestational age. By increasing the resolution of gestational age from weeks (as used in previous studies) to days, we observed nonlinear dependence of FF to gestational age. This may help clinicians to decide whether and when to realize repeat testing.

The best-known factor affecting FF is gestational age. Basically, many studies confirmed that the FF is enhanced with increasing gestational ages. Wang et al. noted that levels of fetal cfDNA increased 0.1% per week between 10 and 21 weeks of gestation, and after 21 weeks of gestation, fetal cfDNA increased 1% per week ^18^. Kinnings et al. showed that the lowest rate of increase in FF with gestational age occurs between 12.5 and 20 weeks gestation (0.083% per week), before increasing tenfold to a rate of 0.821% per week from 20 weeks gestation onwards, which was in general agreement with Wang et al ^24^. Benn et al. demonstrated that the probability that a repeated sampling will provide a result is dependent on the FF at the first draw, maternal weight, and time interval between draws but is not strongly dependent on the gestational age at the time of the first sample. Although FF changes only slowly with gestational age in the late first trimester, most women will show a higher fetal fraction at the second draw ^25^. Our results showed that FF maintained relatively stable before 24 weeks gestation yet increased rapidly thereafter, confirming previous discoveries. Using distribution FF on different days, we found a critical point in 102 days’ gestation (approximately 14 weeks gestation) where we observed a significant decrease in FF. We also found that median FF in 92 days’ gestation corresponds to median FF for the 140 days (Figure 2). Song et al. reported that the FF in the 14 pregnant women also steadily increased with increasing gestational age. However, when individual pregnancies were tracked, two clear differences were observed between gestational time points 11-13 weeks and 15-19 weeks gestation, whereby six women showed an increase in FF and eight women showed a significant decrease. In further investigation of the eight patients with a decrease in FF during this gestation interval from first to second trimester, they found an inverse correlation between FF and maternal weight, indicating that an increase in maternal weight may have been the causative factor ^19^. This is in concordance with our study, as our results also showed a sharp increase in maternal weight in the time interval where FF decreases significantly.

We found that the FF of female fetuses was significantly higher than male fetuses at the same gestational age (Figure 3A), further supported by the differences in their fragment length profiles (Figure 3C). We explained part of this discrepancy with longer female genomes and thus more cfDNA released to the bloodstream in the case of female fetuses. Miltoft et al. reported that the FF decreased significantly with maternal weight and increased significantly with levels of β-hCG and PAPP-A, and among female fetuses in both univariate and multivariate analyses. They reported that FF was about 1% higher in pregnancies with female fetuses ^26^. The biological differences between the sexes, which are apparent even from the early part of the pregnancy, can also explain part of the difference in FF between male and female fetuses. The growth of the male fetuses is greater than the female fetuses, and differences in fetal growth are already present from the first trimester of pregnancy onwards and track throughout pregnancy. Male and female fetuses do not only differ in weight but also differ in biometric indices with a different body proportion as a consequence ^27^. Other differences were identified in placentae of male and female fetuses that have different protein and gene expressions, especially in adverse conditions ^28^. We hypothesize that these biological factors may also contribute to the difference in FF between male and female fetuses.

Our observations of FF between fetal gender and in different gestational ages may have important clinical implications on NIPT timing for pregnant women. Therefore, given that FF can decrease in some pregnant women during a specific time in pregnancy, as demonstrated in this study, we assume that it may be more clinically beneficial to offer NIPT towards between 11-13 weeks gestation where the majority of the women have not substantially gained weight, and the FF is still sufficient to ensure high test sensitivity.

## Data Availability

All data produced in the present study are available upon reasonable request to the authors.

